# Primary care clinical management following self-harm during the first wave of COVID-19 in the UK

**DOI:** 10.1101/2021.03.19.21253969

**Authors:** Sarah Steeg, Matthew J Carr, Laszlo Trefan, Darren M Ashcroft, Nav Kapur, Emma Nielsen, Brian McMillan, Roger T Webb

## Abstract

**Background:** A substantial reduction in GP-recorded self-harm occurred during the first wave of COVID-19 but effects on primary care management of self-harm are unknown.

**Aim:** To examine the impact of COVID-19 on clinical management within three months of an episode of self-harm.

**Design and setting:** Prospective cohort study using data from the UK Clinical Practice Research Datalink.

**Method:** We compared cohorts of patients with an index self-harm episode recorded during a pre-pandemic period (10^th^ March-10^th^ June, 2010-2019) versus the COVID-19 first-wave period (10^th^ March-10^th^ June 2020). Patients were followed up for three months to capture psychotropic medication prescribing, GP/practice nurse consultation and referral to mental health services.

**Results:** 48,739 episodes of self-harm were recorded during the pre-pandemic period and 4,238 during the first-wave COVID-19 period. Similar proportions were prescribed psychotropic medication within 3 months in the pre-pandemic (54.0%) and COVID-19 first-wave (54.9%) cohorts. Likelihood of having at least one GP/practice nurse consultation was broadly similar (83.2% vs. 80.3% in the COVID-19 cohort). The proportion of patients referred to mental health services in the COVID-19 cohort (3.4%) was around half of that in the pre-pandemic cohort (6.5%).

**Conclusion:** Despite the challenges experienced by primary healthcare teams during the initial COVID-19 wave, prescribing and consultation patterns following self-harm were broadly similar to pre-pandemic levels. However, the reduced likelihood of referral to mental health services warrants attention. Accessible outpatient and community services for people who have self-harmed are required as the COVID-19 crisis recedes and the population faces new challenges to mental health.

## Introduction

Prompt clinical intervention and follow-up is recommended for people who have recently self-harmed, in part due to their increased risks of suicide. ^1^ Self-harm includes intentional self-poisoning and self-injury and can involve varying degrees of suicidal intent. ^2^ All episodes of self-harm should be followed by comprehensive mental health assessment to identify psychosocial needs and address risks of further self-harm and suicide. ^1^ This has become more challenging due to the disruption to UK health services caused by the COVID-19 pandemic. Considerable reductions in presentation rates have been reported for a number of physical and mental health conditions, ^3^ including self-harm. ^4^ The pandemic and its pervasive impacts on everyday life has also had a detrimental impact on the mental health of the population ^5^ and the first wave of COVID-19 and lockdown may have led to an increase in the prevalence of suicidal ideation. ^6^ This unique combination of factors has led to a shortfall in the numbers of people receiving support from healthcare services after harming themselves.

In April 2020, during the first UK COVID-19 lockdown, rates of primary care-recorded incident self-harm in the UK were 38% lower than expected based on trends that occurred during the previous ten years. ^4^ While rates of help seeking gradually returned towards expected levels through the subsequent months up to September 2020, further regional COVID-19 containment restrictions and national lockdowns from autumn 2020 into winter 2021 are likely to have additionally affected rates of help seeking. Some people sought help from alternative sources whilst the UK was in its first lockdown; for example, some mental health charities reported increases in demand for services such as helplines. ^7^ One study in the US reported emergency hospital presentations following suicide attempts had increased to higher than pre-pandemic levels following an initial reduction, ^8^ suggesting that observed reductions in help seeking in primary care did not necessarily reflect population need.

Primary care settings provide vital support for people who have self-harmed. A recent study found that 26% of people sought help from their general practitioner (GP) in the week prior to suicide, with self-harm a common reason for contact.^9^ Previous research found that 15% of patients with an episode of self-harm recorded in primary care were referred to mental health services from their GP within a year. ^10^ A recent report found that, among people referred to mental health services, their GP was the most common referral route. ^11^ However, this report highlighted many barriers that exist for people accessing support following self-harm. ^11^ GPs, as well as patients, have reported struggling to find appropriate self-harm services, with limited referral options and shortages in community services identified. ^12^

Although GPs continued to offer consultations to patients throughout the first wave in the spring of 2020, with most consultations taking place remotely, ^13^ recently conducted research found lower rates of GP referral to mental health services following primary care-recorded common mental illnesses and self-harm episodes. ^4^ The degree to which primary care management of people who had self-harmed was impacted during the first COVID-19 wave is unknown. In the UK, a nationwide lockdown was imposed on 23^rd^ March 2020, with public health messaging to encourage people to avoid contact with others announced the week before this. ^15^ We aimed to examine clinical management of self-harm during the three months following the beginning of the UK COVID-19 containment measures and national lockdown, using data from a pre-pandemic comparison period to examine effects of COVID-19. Our specific objectives were to:

i. Identify two cohorts of patients presenting with an index episode of self-harm, comprising pre-pandemic versus COVID-19 first-wave time periods.
ii. Estimate the likelihood of receiving a new psychotropic medication prescription (by drug type), frequency of subsequent GP or practice nurse consultations, and referral to mental health services within 3 months of the index self-harm episode.
iii. Estimate rate ratios for frequency of psychotropic medication prescribing, referral to mental health services and GP or practice nurse consultation (by face-to-face and telephone) within three months of the index episode, between the COVID-19 and pre-pandemic comparison cohorts.

## Methods

### Study design, data sources, and participants

We conducted a prospective cohort study using anonymised primary care data from the Clinical Practice Research Datalink (CPRD) Aurum and GOLD databases. ^16, 17^ The Aurum database includes general practices based in England that contribute data using the EMIS® clinical system. The GOLD database is extracted from the Vision® system, with most of its contributing general practices based in Northern Ireland, Scotland and Wales. To avoid including duplicate general practices in the Aurum and GOLD databases, we excluded English practices in our analyses of GOLD data. The CPRD includes information on patient demographics, consultations, symptoms, diagnoses, medication prescriptions and referrals to secondary care. We also obtained information on IMD linked at general practice level. ^18^ The IMD is a composite measure of the relative deprivation of an area derived from several domains including income, employment and health. ^19^

Analysis was conducted on pooled Aurum and GOLD data. We compared two cohorts of patients: (i) those with an index self-harm episode recorded in a pre-pandemic comparison period (between 10^th^ March to 10^th^ June, 2010-2019) and those with an index self-harm episode recorded in the COVID-19 first-wave period (10^th^ March to 10^th^ June 2020). Patients in each cohort were followed up for three months to capture psychotropic medication prescribing, referral to mental health services (available in GOLD only) and GP or practice nurse consultations. To be included in either of the two study cohorts, patients must have been aged 10 years or older and registered with a contributing practice, deemed by the CPRD as providing up-to-standard data, for at least one year prior to the date of the index self-harm episode. Patients with less than 3 months of follow-up time in the CPRD were excluded from our analyses. The cohorts were restricted to patients with records that were deemed acceptable by the CPRD for research purposes, which excluded patients with missing data on sex or age. IMD data were missing for 9.2% of the pre-pandemic comparison cohort and 10.3% of the COVID-19 first-wave cohort (Table 1).

**Table 1.** Characteristics of cohorts with a primary care-recorded episode of self-harm in the UK

|  |  |  |
| --- | --- | --- |
|  | Comparison cohort<br>(2010-2019) | COVID-19 cohort<br>(2020) |
| Total index episodes<br>(between 10<sup>th</sup> March<br>and 10<sup>th</sup> June) | 48,739 | 4,238 |
| Gender: |  |  |
| Female | 29,596 (60.7%) | 2,510 (59.2%) |
| Male | 19,143 (39.3%) | 1,728 (40.8%) |
| Age group (yrs): |  |  |
| 10-24 | 20,308 (41.7%) | 1,844 (43.5%) |
| 25-64 | 25,700 (52.7%) | 2,159 (50.9%) |
| ≥65 | 2,731 (5.6%) | 235 (5.6%) |
| Practice-level IMD quintile: |  |  |
| 1 (least deprived) | 5,687 (11.7%) | 510 (12.0%) |
| 2 | 7,003 (14.4%) | 600 (14.2%) |
| 3 | 8,035 (16.5%) | 729 (17.2%) |
| 4 | 10,837 (22.2%) | 980 (23.1%) |
| 5 (most deprived) | 12,678 (26.0%) | 981 (23.2%) |
| Unknown | 4,499 (9.2%) | 438 (10.3%) |

The study was approved by the Independent Scientific Advisory Committee (protocol number 20_001RA2).

### Exposures, outcomes and covariates

Index episodes of self-harm were identified from Read, SNOMED and EMIS codes. To identify individuals’ first record of self-harm, we applied a retrospective analysis period during which a patient was required to have been registered at the practice for at least 1 year before an incident episode. We examined any psychotropic medication and specific psychotropic medication types (https://clinicalcodes.rss.mhs.man.ac.uk/medcodes/article/173/). Mode of consultation was grouped into face-to-face and video/telephone consultations, with a GP or practice nurse. Information on referral to mental health services was only available in the GOLD data. Two fields were used to identify referrals: a ‘psychiatry’ code in the Family Health Services Authority variable and codes of ‘mental illness’, ‘child and adolescent psychiatry’, ‘forensic psychiatry’, ‘psychotherapy’, ‘old age psychiatry’, ‘clinical psychology’, ‘adult psychiatry’ and ‘community psychiatric nurse’ in the National Health Service specialty field. ^10^ We combined information from both the FHSA and NHS fields to construct a binary specialist mental health services referral indicator. All code lists were verified by senior clinical academics as part of a previous study ^4^ and are available online (https://clinicalcodes.rss.mhs.man.ac.uk/medcodes/article/173/). We examined frequencies and rate ratios between the pre-pandemic and COVID-19 first-wave cohorts by gender, age group (10-24, 25-64 and 65 and older) and practice-level IMD quintile.

### Analysis

Frequencies of psychotropic medication prescribing, GP/nurse consultation and referral to mental health services during the antecedent period were estimated and compared to observed values during the COVID-19 first-wave period. The modelling was conducted using modified Poisson regression in a generalised linear modelling framework with a log-link function and a robust variance estimator with analyses stratified by gender, age group and practice-level IMD quintile.

This study was conducted in accordance with REporting of studies Conducted using Observational Routinely-collected health Data guidance (RECORD; Table S1). ^20^ A panel of four service users and carers with lived experience of health services following self-harm collaborated with the research team to plan the study and interpret results.

## Results

A total of 48,739 patients had an index episode of self-harm recorded in England during the pre-pandemic comparison period and 4,238 were recorded in the COVID-19 first-wave cohort (Table 1). The gender, age and deprivation profiles of the two cohorts were broadly similar, with the majority of recorded self-harm episodes by women and more self-harm episodes recorded in practices in areas of higher deprivation.

Just over half of patients in the pre-pandemic (54.0%) and COVID-19 first-wave (54.9%) cohorts were prescribed psychotropic medication within three months of the index self-harm episode (Table 2). The likelihoods of receiving such treatment were broadly similar across gender and deprivation quintiles. Among patients aged 10-24 years, those in the COVID-19 first-wave cohort were more likely to be prescribed psychotropic medication (ratio 1.14, 95% CI 1.07 to 1.22). Considering prescriptions for antidepressant medication specifically, probabilities were broadly similar in the pre-pandemic and COVID-19 first-wave cohorts, though they were higher among young people aged 10-24 years in the COVID-19 first-wave cohort (ratio 1.18, CI 1.10 to 1.26). Details on prescribing of antipsychotic, anxiolytic/hypnotic, mood stabilisers and stimulants are in supplementary Table S2.

**Table 2.**
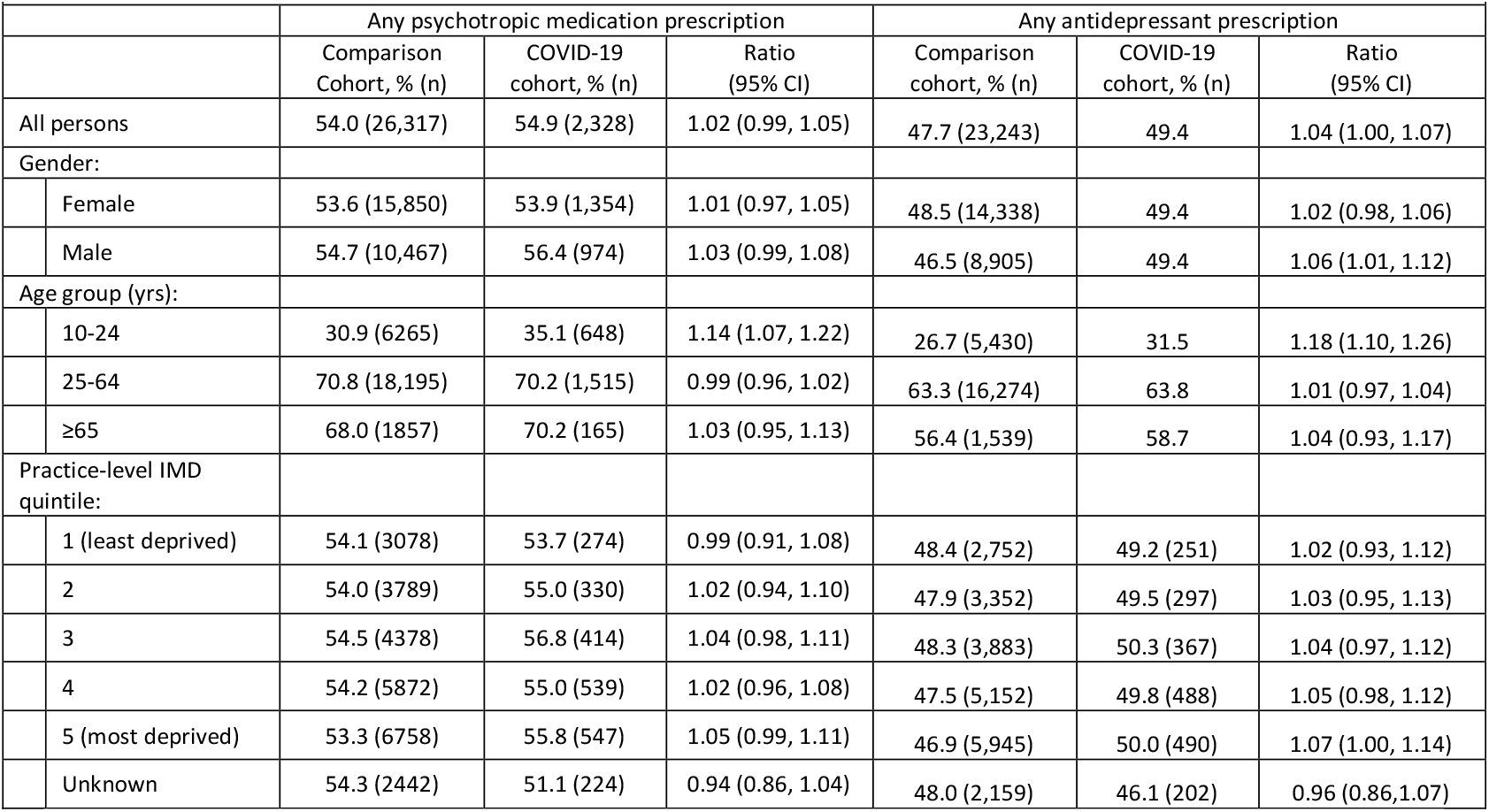
Prescriptions of any type of psychotropic medication and any antidepressant drug following a primary care-recorded episode of self-harm in the UK

Unsurprisingly, the likelihood of patients receiving a remote GP/practice nurse consultation within 3 months of a self-harm episode was higher in the COVID-19 ‘first-wave’ cohort (67.7%) than in the pre-pandemic comparison cohort (32.3%, ratio 2.10, CI 2.05 to 2.15) (Table 3). Although the overall likelihood of having a GP/practice nurse consultation was slightly lower in the COVID-19 cohort (80.3%) than in the pre-pandemic comparison cohort (83.2%), ratio 0.97, CI 0.96 to 0.98, this pattern did not apply to all demographic groups. Men, patients aged 65 years and over and those registered with practices in the two most deprived quintiles were equally likely to have had a GP/practice nurse consultation in the pre-pandemic and COVID-19 first-wave cohorts.

**Table 3.** GP/practice nurse consultations following a primary care-recorded episode of self-harm in the UK

|  |  |  |  |  |  |  |  |  |  |  |
| --- | --- | --- | --- | --- | --- | --- | --- | --- | --- | --- |
|  |  | Face-to-face consultation |  |  | Telephone/Video consultation |  |  | Face-to-Face or Telephone/Video consultation |  |  |
|  |  | Comparison cohort, % (n) | COVID-19 cohort, % (n) | Ratio (95% CI) | Comparison cohort, % (n) | COVID-19 cohort, % (n) | Ratio (95% CI) | Comparison cohort, % (n) | COVID-19 cohort, % (n) | Ratio (95% CI) |
| All persons |  | 80.1 (39,027) | 54.3 (2,300) | 0.68 (0.66, 0.70) | 32.3 (15,723) | 67.6 (2,866) | 2.10 (2.05, 2.15) | 83.2 (40,536) | 80.3 (3,403) | 0.97 (0.95, 0.98) |
| Gender: |  |  |  |  |  |  |  |  |  |  |
|  | Female | 82.5 (24,423) | 56.3 (1,412) | 0.68 (0.66, 0.71) | 33.8 (10,004) | 69.8 (1,752) | 2.06 (2.00, 2.13) | 85.5 (25,300) | 82.1 (2,060) | 0.96 (0.94, 0.98) |
|  | Male | 76.3 (14,604) | 51.4 (888) | 0.67 (0.64, 0.71) | 29.9 (5,719) | 64.5 (1,114) | 2.16 (2.07, 2.25) | 79.6 (15,236) | 77.7 (1,343) | 0.98 (0.95, 1.00) |
| Age group (yrs): |  |  |  |  |  |  |  |  |  |  |
|  | 10-24 | 74.3 (15,083) | 48.2 (888) | 0.65 (0.62, 0.68) | 24.9 (5,055) | 60.5 (1,116) | 2.43 (2.33, 2.54) | 77.4 (15,720) | 74.7 (1,377) | 0.96 (0.94, 0.99) |
|  | 25-64 | 84.0 (21,596) | 58.0 (1,252) | 0.69 (0.67, 0.72) | 36.4 (9,341) | 72.8 (1,571) | 2.00 (1.94, 2.06) | 86.9 (22,327) | 84.0 (1,813) | 0.97 (0.95, 0.99) |
|  | ≥65 | 86.0 (2,348) | 68.1 (160) | 0.79 (0.72, 0.87) | 48.6 (1,327) | 76.2 (179) | 1.57 (1.45, 1.70) | 91.1 (2,489) | 90.6 (213) | 0.99 (0.95, 1.04) |
| Practice-level IMD quintile: |  |  |  |  |  |  |  |  |  |  |
|  | 1 (least deprived) | 82.4 (4,687) | 56.9 (290) | 0.69 (0.64, 0.74) | 37.5 (2,135) | 68.0 (347) | 1.81 (1.69, 1.94) | 86.0 (4,891) | 81.0 (413) | 0.94 (0.90, 0.98) |
|  | 2 | 81.6 (5,711) | 50.8 (305) | 0.62 (0.58, 0.67) | 33.4 (2,340) | 70.2 (421) | 2.10 (1.97, 2.23) | 84.5 (5,918) | 80.0 (480) | 0.95 (0.91, 0.99) |
|  | 3 | 81.3 (6,534) | 55.8 (407) | 0.69 (0.64, 0.73) | 34.0 (2,731) | 66.3 (483) | 1.95 (1.84, 2.07) | 84.7 (6,807) | 80.0 (583) | 0.94 (0.91, 0.98) |
|  | 4 | 79.5 (8,613) | 53.7 (526) | 0.68 (0.64, 0.72) | 32.0 (3,472) | 67.5 (661) | 2.11 (2.00, 2.22) | 82.6 (8,949) | 80.8 (792) | 0.98 (0.95, 1.01) |
|  | 5 (most deprived) | 79.1 (10,023) | 56.2 (551) | 0.71 (0.67, 0.75) | 29.3 (3,717) | 68.3 (670) | 2.33 (2.21, 2.45) | 81.7 (10,358) | 81.9 (803) | 1.00 (0.97, 1.03) |
|  | Unknown | 76.9 (3,459) | 50.5 (221) | 0.66 (0.60, 0.72) | 29.5 (1,328) | 64.8 (284) | 2.20 (2.02, 2.39) | 80.3 (3,613) | 75.8 (332) | 0.94 (0.89, 1.00) |

Information on primary care referrals to mental health services was only available in the GOLD dataset (representing practices in Northern Ireland, Scotland and Wales). Overall, 3.4% of patients (31/910) were referred to mental health services in the COVID-19 first-wave cohort, around half of that in the pre-pandemic cohort (6.5%, ratio 0.53, CI 0.37 to 0.75) (Table 4). The reduction in likelihood of being referred to mental health servicers was greater among women (ratio 0.34, CI 0.19 to 0.60) than for men (0.83, CI 0.53 to 1.30), p value for effect modification by gender = 0.016). The fall in probability of being referred also appeared to be greater for patients aged 10-24 years (ratio 0.33, CI 0.17 to 0.66). However, the numbers of patients following stratification by referral status and age group was not sufficiently powered to conduct formal tests for interaction effects in these groups.

**Table 4.**
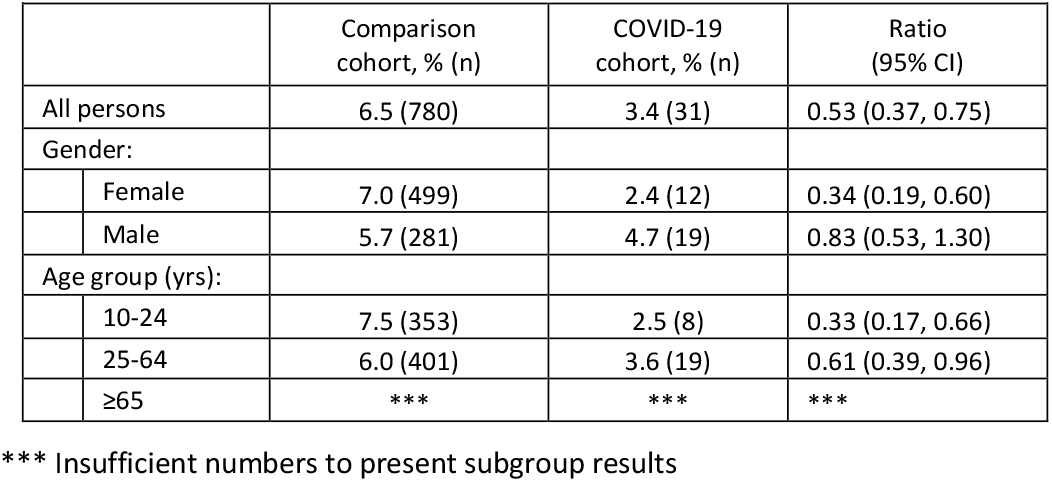
Referrals to mental health services following a primary care-recorded episode of self-harm in Northern Ireland, Scotland and Wales (practices from CPRD GOLD)

## Discussion

### Summary

Pre-pandemic and COVID-19 first-wave cohorts of patients with an index episode of self-harm recorded in primary care had similar gender, age and deprivation profiles. Similar proportions were prescribed psychotropic medication within three months of their index self-harm episode - just over half in both the pre-pandemic and COVID-19 first-wave cohorts. However, patients aged 10-24 years in the COVID-19 first-wave cohort were more likely to be prescribed psychotropic medication than in the preceding years. Overall, the likelihood of having at least one GP/practice nurse consultation was slightly lower in the COVID-19 first-wave cohort, although there was no such difference observed among men, patients aged 65 years and over, and those registered with practices located in more deprived areas. The proportion of patients referred to mental health services (in Northern Ireland, Scotland and Wales) in the COVID-19 first-wave cohort was around half of that in the pre-pandemic comparison cohort with larger deficits in referral likelihood observed among women.

### Strengths and limitations

The main strength of our study is the broadly representative data source that included a large number of general practices across the UK. CPRD Aurum is broadly representative of geographical coverage, area-level deprivation, age, and sex in England ^16^ while CPRD GOLD dataset is broadly representative of the UK age and sex profile.^17^ This enables us to make inferences at national level about how the pandemic affected primary care clinical management of patients who have self-harmed. However, there are some limitations in utilising anonymised primary care records. The data are extracted from GP information systems and their accuracy is determined by the quality of the information inputted by contributing practices. Some of the self-harm episodes recorded in primary care would have been emergency department presentations that were subsequently added to the patient’s primary care record. Future research using linked Hospital Episode Statistics will enable separate examination of emergency department self-harm presentations. Finally, we were unable to examine referrals to mental health services in England due to limitations to the CPRD Aurum dataset.

### Comparison with existing literature

Previous research found that reductions in help-seeking were greatest among patients registered at practices located in more deprived areas.^4^ Our study found that patients at practices in the two most deprived quintiles who did seek help had similar rates of psychotropic medication prescribing as those at practices in less deprived areas, in both the pre-pandemic comparison and COVID-19 first-wave cohorts. Similarly, the abrupt switch to remote consultations once the COVID-19 crisis had commenced occurred to a similar degree at practices across the practice population deprivation spectrum. This suggests that for people who did seek help, there was no exacerbation of existing inequalities due to COVD-19 in these particular aspects of primary care clinical management.

However, likelihood of referral to mental health services was lower during the first wave of COVID-19 than during the pre-pandemic comparison period, particularly for women. Younger people may also have been less likely to receive a referral for mental health services (though our findings lacked statistical power to infer this with certainty). Both of these groups have been found to have been particularly negatively affected by the pandemic. Individuals aged 18-29 years reported greater increases in suicidal ideation over the first six weeks of the UK’s lockdown than other groups, ^6^ while women and younger people were found to have the greatest deteriorations in mental health. ^5^ Furthermore, these groups were previously identified as having the greatest reductions in help seeking for mental illness and self-harm during April 2020.^4^

### Implications for research and practice

Evidence shows the number of people seeking mental health help, including for self-harm, from non-NHS services such as via digital platforms and helplines increased during the second quarter of 2020. ^21, 22^ This has implications for the clinical guidance for people who have self-harmed, which recommends that assessment by a mental health specialist should follow all episodes of self-harm. ^14^ Furthermore, such assessments might be particularly challenging in non-face-to-face settings.

Data from NHS Digital ^23^ showed a 10% decrease in the number of new referrals to NHS mental health services in the six months from 1^st^ April 2020. During the same time period, there was an increase in antidepressant prescribing of around 4% leading to concerns that the increased mental health burden caused by the COVID-19 crisis could be being managed pharmacologically rather than with psychosocial interventions. Our findings suggest this could also have been happening with patients who have harmed themselves. Whilst outpatient and community mental health services adapted to provide alternatives to face-to-face support, our findings suggest that primary care practitioners were less inclined to refer to these services during the early stages of the pandemic. Ongoing work in England to improve community support following self-harm emphasises a need to better align the voluntary sector with primary healthcare services, ^24^ to ensure that GPs are equipped with the information that they require to make referrals to community self-harm resources as well as NHS services.

## Conclusions

Despite the challenges experienced by general practitioners in delivering health care during the three months of the initial wave of COVID-19 in the UK, the management of self-harm was broadly similar to the pre-pandemic comparison period in terms of psychotropic prescribing and GP/nurse consultations. However, the reduced likelihood of referral to mental health services during March-June 2020 warrants close attention, particularly in the context of increasing prevalence of mental distress in the population and especially so for women. Our findings suggest that COVID-19 may have increased the likelihood that adolescents and young people do not receive psychosocial interventions, with pharmacological intervention alone becoming more likely during the early phase of the crisis. Accessible outpatient and community services that people who have self-harmed can be referred to by their GPs are required.

## Data Availability

The clinical codes used in this study are available online. The codes are also available from the corresponding author on request. Access to data are available only once approval has been obtained through the individual constituent entities controlling access to the data. The primary care data can be requested via application to the Clinical Practice Research Datalink.

## Author contributions

All authors conceptualised the study and contributed to its design. MJC, LT and SS accessed and verified the data. MJC generated the clinical code lists. MJC and LT managed the data, did statistical analysis. SS drafted the manuscript. All authors critically reviewed the manuscript and approved the final version.

## Ethical approval

This study was done using data from the CPRD obtained under licence from the UK Medicines and Healthcare products Regulatory Agency (MHRA). The data are provided by patients and collected by the NHS as part of their care and support. All patient data were de-identified; thus, the requirement for patient consent was waived. Individual patients can opt out of sharing their records with the CPRD. The study protocol was approved by the Independent Scientific Advisory Committee (protocol reference number 20_001R2).

## Acknowledgements

We thank Stephen Barlow, Elizabeth Monaghan, Fiona Naylor, and Jonathan Smith (members of the Centre for Mental Health and Safety patient and public involvement and engagement group of the NIHR Greater Manchester Patient Safety Translational Research Centre) for their advisory roles in the study. We would also like to acknowledge all the data providers and general practices that made the anonymised data available for research.

## Supplementary material

**Table S1:** The RECORD statement – checklist of items, extended from the STROBE statement, that should be reported in observational studies using routinely collected health data.

|  | Item No. | STROBE items | Location in manuscript where items are reported | RECORD items | Location in manuscript where items are reported |
| --- | --- | --- | --- | --- | --- |
| **Title and abstract** |  |  |  |  |  |
|  | 1 | (a) Indicate the study's design with a commonly used term in the title or the abstract (b) Provide in the abstract an informative and balanced summary of what was done and what was found |  | RECORD 1.1: The type of data used should be specified in the title or abstract. When possible, the name of the databases used should be included.<br>RECORD 1.2: If applicable, the geographic region and timeframe within which the study took place should be reported in the title or abstract.<br>RECORD 1.3: If linkage between databases was conducted for the study, this should be clearly stated in the title or abstract. | Abstract: 'Design and setting'.<br>Abstract: 'Method'. |
| **Introduction** |  |  |  |  |  |
| Background rationale | 2 | Explain the scientific background and rationale for the investigation being reported |  |  | Introduction: paragraphs 1-5 |
| Objectives | 3 | State specific objectives, including any prespecified hypotheses |  |  | Introduction: paragraph 5 |
| **Methods** |  |  |  |  |  |
| Study Design | 4 | Present key elements of study design early in the paper |  |  | Methods: 'Study design, data sources, and participants' |
| Setting | 5 | Describe the setting, locations, and relevant dates, including periods of recruitment, exposure, follow-up, and data collection |  |  | Methods: 'Study design, data sources, and participants' |
| Participants | 6 | (a) *Cohort study* - Give the eligibility criteria, and the sources and methods of selection of participants. Describe methods of follow-up<br>*Case-control study* - Give the eligibility criteria, and the sources and methods of case ascertainment and control selection. Give the rationale for the choice of cases and controls<br>*Cross-sectional study* - Give the eligibility criteria, and the sources and methods of selection of participants<br>(b) *Cohort study* - For matched studies, give matching criteria and number of exposed and unexposed<br>*Case-control study* - For matched studies, give matching criteria and the number of controls per case |  | RECORD 6.1: The methods of study population selection (such as codes or algorithms used to identify subjects) should be listed in detail. If this is not possible, an explanation should be provided.<br>RECORD 6.2: Any validation studies of the codes or algorithms used to select the population should be referenced. If validation was conducted for this study and not published elsewhere, detailed methods and results should be provided.<br>RECORD 6.3: If the study involved linkage of databases, consider use of a flow diagram or other graphical display to demonstrate the data linkage process, including the number of individuals with linked data at each stage. | Methods: ‘*Study design, data sources, and participants*’ (paragraph 3) and ‘*Exposures, outcomes and covariates*’<br>Results: paragraph 1 |
| Variables | 7 | Clearly define all outcomes, exposures, predictors, potential confounders, and effect modifiers. Give diagnostic criteria, if applicable. |  | RECORD 7.1: A complete list of codes and algorithms used to classify exposures, outcomes, confounders, and effect modifiers should be provided. If these cannot be reported, an explanation should be provided. | Methods: ‘*Exposures, outcomes and covariates*’ |
| Data sources/<br>measurement | 8 | For each variable of interest, give sources of data and details of methods of assessment (measurement). Describe comparability of assessment methods if there is more than one group |  |  | Methods: ‘*Exposures, outcomes and covariates*’ |
| Bias | 9 | Describe any efforts to address potential sources of bias |  |  | Methods: ‘*Analysis*’ |
| Study size | 10 | Explain how the study size was arrived at |  |  | Methods: 'Study design, data sources, and participants' (paragraph 2) |
| Quantitative variables | 11 | Explain how quantitative variables were handled in the analyses. If applicable, describe which groupings were chosen, and why |  |  | Methods: 'Study design' |
| Statistical methods | 12 | (a) Describe all statistical methods, including those used to control for confounding<br>(b) Describe any methods used to examine subgroups and interactions<br>(c) Explain how missing data were addressed<br>(d) *Cohort study* - If applicable, explain how loss to follow-up was addressed<br>*Case-control study* - If applicable, explain how matching of cases and controls was addressed<br>*Cross-sectional study* - If applicable, describe analytical methods taking account of sampling strategy<br>(e) Describe any sensitivity analyses |  |  | Methods: 'Analysis' |
| Data access and cleaning methods |  | .. |  | RECORD 12.1: Authors should describe the extent to which the investigators had access to the database population used to create the study population.<br>RECORD 12.2: Authors should provide information on the data cleaning methods used in the study. | 'Author contributions', page 12 |
| Linkage |  | .. |  | RECORD 12.3: State whether the study included person-level, institutional-level, or other data linkage across two or more databases. The methods of linkage and methods of linkage quality evaluation should be provided. | Methods: 'Data sources' and 'Data analysis' |
| **Results** |  |  |  |  |  |
| Participants | 13 | (a) Report the numbers of individuals at each stage of the study (*e.g.*, numbers potentially eligible, examined for eligibility, confirmed eligible, included in the study, completing follow-up, and analysed)<br>(b) Give reasons for non-participation at each stage.<br>(c) Consider use of a flow diagram |  | RECORD 13.1: Describe in detail the selection of the persons included in the study (*i.e.*, study population selection) including filtering based on data quality, data availability and linkage. The selection of included persons can be described in the text and/or by means of the study flow diagram. | Results: paragraph 1 |
| Descriptive data | 14 | (a) Give characteristics of study participants (*e.g.*, demographic, clinical, social) and information on exposures and potential confounders<br>(b) Indicate the number of participants with missing data for each variable of interest<br>(c) *Cohort study* - summarise follow-up time (*e.g.*, average and total amount) |  |  | Results: paragraph 1<br>Methods: '*Study design, data sources, and participants*' (paragraph 2)<br>Methods: '*Study design, data sources, and participants*' (paragraph 2) |
| Outcome data | 15 | *Cohort study* - Report numbers of outcome events or summary measures over time<br>*Case-control study* - Report numbers in each exposure category, or summary measures of exposure<br>*Cross-sectional study* - Report numbers of outcome events or summary measures |  |  | Table 1 |
| Main results | 16 | (a) Give unadjusted estimates and, if applicable, confounder-adjusted estimates and their precision (e.g., 95% confidence interval). Make clear which confounders were adjusted for and why they were included<br>(b) Report category boundaries when continuous variables were categorized<br>(c) If relevant, consider translating estimates of relative risk into absolute risk for a meaningful time period |  |  | Results and Tables 1-4. |
| Other analyses | 17 | Report other analyses done—e.g., analyses of subgroups and interactions, and sensitivity analyses |  |  |  |
| **Discussion** |  |  |  |  |  |
| Key results | 18 | Summarise key results with reference to study objectives |  |  | Discussion: 'Summary'. |
| Limitations | 19 | Discuss limitations of the study, taking into account sources of potential bias or imprecision. Discuss both direction and magnitude of any potential bias |  | RECORD 19.1: Discuss the implications of using data that were not created or collected to answer the specific research question(s). Include discussion of misclassification bias, unmeasured confounding, missing data, and changing eligibility over time, as they pertain to the study being reported. | Discussion: 'Strengths and limitations' |
| Interpretation | 20 | Give a cautious overall interpretation of results considering objectives, limitations, multiplicity of analyses, results from similar studies, and other relevant evidence |  |  | Discussion: 'Implications for research and practice' |
| Generalisability | 21 | Discuss the generalisability (external validity) of the study results |  |  | Discussion: 'Strengths and limitations' |
| **Other Information** |  |  |  |  |  |
| Funding | 22 | Give the source of funding and the role of the funders for the present study and, if applicable, for the original study on which the present article is based |  |  | Funding statement |
| Accessibility of protocol, raw data, and programming code |  | .. |  | RECORD 22.1: Authors should provide information on how to access any supplemental information such as the study protocol, raw data, or programming code. | Data sharing statement |

\*Reference: Benchimol EI, Smeeth L, Guttman A, Harron K, Moher D, Petersen I, Sørensen HT, von Elm E, Langan SM, the RECORD Working Committee. The REporting of studies Conducted using Observational Routinely-collected health Data (RECORD) Statement. *PLoS Medicine* 2015; in press.
\*Checklist is protected under Creative Commons Attribution ([CC BY](https://creativecommons.org/licenses/by/4.0/)) license.

**Table S2.**
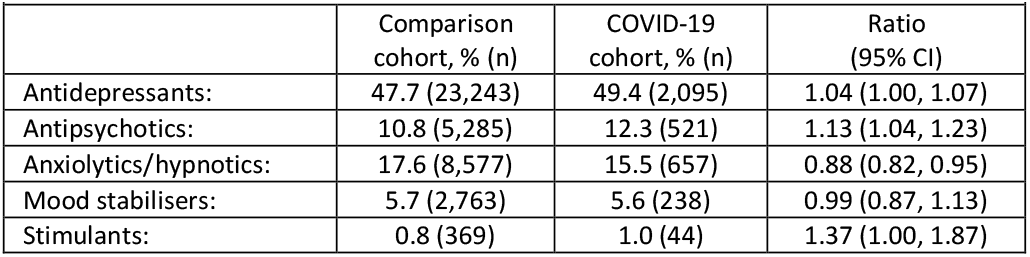
Prescriptions of specific psychotropic medication types following a primary care-recorded episode of self-harm in the UK

